# Percutaneous Auricular Nerve Stimulation for Treating Post-COVID Fatigue (PAuSing-pCF): a single-site, single-blind, sham-controlled, randomized, interventional, crossover study protocol

**DOI:** 10.1101/2025.07.31.25332440

**Authors:** Maria Germann, Svetlana Cherlin, Marzieh Shahmandi, Aidan S. Baker, Fai Ng, Demetris S. Soteropoulos, Stuart N. Baker, James M. S. Wason, Mark R. Baker

## Abstract

**Background:** According to the Office for National Statistics, an estimated 2.3% of people in the UK suffer from post-COVID Fatigue (pCF) following acute COVID-19 infection. Feedback from those with pCF has highlighted the devastating impact it has on their lives and the need for novel therapeutic options. Fatigue encompasses not only the perception of increased physical effort and extreme tiredness but also cognitive/mental fatigue. As part of a previous study (Baker et al., 2023), we recently showed that fatigue in pCF correlates with dysregulation in specific components of the central, peripheral and autonomic nervous systems. The vagus nerve is a major component of the autonomic nervous system and plays an important role both in metabolic homeostasis and in inflammatory modulation. Transcutaneous auricular vagus nerve stimulation (taVNS) offers an easy and reliable approach to activating the vagus nerve non-invasively and can be self-administered safely at home. This study will probe the mechanisms of pCF in adults by testing whether taVNS self-administered using a transcutaneous electrical nerve stimulation (TENS) device can reduce symptoms of fatigue and normalise changes in the peripheral and central nervous system that are hypothesized to mediate fatigue.

**Design:** This will be a single-site, single-blind, sham-controlled, randomized, interventional trial in 96 people with pCF. Participants will be randomized to one of three groups, one active and two control interventions, which they will self-administer for 8 weeks. After 8 weeks everyone will crossover to the active intervention for another 8 weeks of taVNS. Ongoing levels of fatigue will be assessed using visual analogue scales and a battery of relevant questionnaires. All participants will undergo extensive electrophysiological testing at week 0, week 8 and week 16. Ambulatory data (heart rate variability, activity levels) will be collected using wearable technology.

**Discussion:** This study will establish whether vagus nerve dysfunction is important mechanistically in pCF and whether boosting vagus nerve activity by taVNS can not only improve symptoms of fatigue in pCF but normalise biological and behavioural correlates of fatigue in otherwise healthy members of the public with pCF.

**Trial registration:** This trial is prospectively registered at www.isrctn.com/ISRCTN18015802. Registered May 12, 2022.

## Background

Long COVID has emerged as an unfortunate and common complication of the Corona Virus Disease 2019 (COVID-19) pandemic, affecting both those with severe disease requiring admission to hospital and those with relatively mild acute symptoms of COVID-19 infection. An estimated 3.1% of the UK population were experiencing long COVID symptoms (Office for National Statistics, 2023). Of the many symptoms of Long COVID, or post-acute sequelae of SARS-CoV-2 infection, the commonest is fatigue; post-COVID fatigue (pCF) affects 71% of those with Long COVID in the UK (Office for National Statistics, 2023) which amounts to 2.3% of people in the UK suffering from acute COVID-19 infection.

Whilst the underlying processes that lead to symptomatic fatigue in post-viral fatigue, chronic fatigue syndrome (CFS), fatigue associated with chronic autoimmune diseases, etc. share common mechanisms, these are undoubtedly multi-factorial. However, immune dysregulation is one shared feature frequently encountered in patients with fatigue, regardless of aetiology. For example, antagonising the effects of pro-inflammatory cytokines can improve fatigue in those with autoimmune disease (Tarn et al., 2019). Reduced monoaminergic neurotransmission and neuromodulation have also been implicated in fatigue associated with chronic inflammatory diseases. Several mechanisms appear to be at play and include the action of cytokines (reducing neurotransmitter release at synaptic terminals increasing uptake and breakdown of serotonin, noradrenaline and dopamine) and the effects of chronic inflammation and oxidative stress (reducing monoaminergic neurotransmission and biosynthesis of serotonin; Korte and Straub (2019)).

Fatigue encompasses not only the perception of increased physical effort and extreme tiredness but also cognitive/mental fatigue (problems with thinking, remembering and concentrating). These symptoms must emerge through changes at multiple levels of the peripheral and central nervous system. We previously hypothesized several plausible mechanisms by which immune or neuromodulator dysfunction could lead to changes in the nervous system and cause fatigue (Baker et al., 2023). This study tested 35 physiological, neurophysiological and behavioural measures in individuals with pCF compared to an age-matched control population without pCF and showed that 10 measures are abnormal. These relate to three broad areas: dysautonomia, cortical excitability and muscle pathology. Interestingly, most of these results could be explained by the failure of monoaminergic and/or cholinergic neuromodulation. For example, the prefrontal cortex, which partially mediates response inhibition (Guo et al., 2018) and thus Stop-signal reaction time (SSRT; one of the cortical behavioural measures), is an important node in the cortical-subcortical network of cognitive/mental fatigue (Wylie et al., 2020); prefrontal cortex is modulated by monoaminergic and cholinergic inputs from brainstem and basal forebrain nuclei (Balachandran et al., 2018). Post-infectious impairments in such systems could thus cause cognitive/mental fatigue and affect the SSRT.

These observations are important in formulating hypotheses regarding the mechanisms of post-viral fatigue, specifically pCF; however, they are only correlations. For example, epiphenomena, including adaptive responses to pCF, will also show correlation but are irrelevant mechanistically. Although statistical proof of causality is difficult, Bradford-Hill proposed a number of criteria, one of which is the observation of significant change in correlation when a putative causal factor is perturbed (Bradford, 1965).

Whilst the severe acute respiratory syndrome coronavirus (SARS-CoV-2), which causes COVID-19 is not primarily neurotropic, neurological complications, mediated by secondary coagulopathies or immune-mediated phenomena, are common (Varatharaj et al., 2020, Spudich and Nath, 2022). Such neurological complications include isolated or multiple cranial neuropathies, and of these, vagus neuropathy is also recognized (Moyano et al., 2021) and may be a predictor of severe COVID-19 (Mol et al., 2021). Given the vagus nerve modulates immune processes via the cholinergic anti-inflammatory reflex (Tracey, 2002, Koopman et al., 2016) and that stimulation of the vagus nerve can activate central adrenergic/serotonergic (Hulsey et al., 2019) and cholinergic (Hulsey et al., 2016) pathways, it could be hypothesized that pCF is mediated by vagus nerve dysfunction. By stimulating the vagus nerve in individuals with pCF and measuring changes in neurophysiological and behavioural measures, inflammatory markers, and symptoms of fatigue, it should be possible to test this hypothesis and have a clearer idea as to which biological changes in pCF are cause or effect. Such an approach may also provide evidence for targeting the vagus nerve in managing the symptoms of fatigue in pCF.

Evidence that this approach might work comes from studies in patients with fatigue associated with primary Sjögren’s syndrome (a chronic autoimmune disorder), which showed rapid reductions in fatigue after 4-5 weeks of daily vagus nerve stimulation (Tarn et al., 2019). Similar positive findings were also reported in patients with fatigue associated with another chronic autoimmune disorder, systemic lupus erythematosus (Aranow et al., 2021).

Vagus nerve stimulation (VNS) is a well-established therapy (e.g. for intractable epilepsy), requiring surgical implantation of electrodes around the vagus nerve in the neck and a stimulator subcutaneously in the chest. In epilepsy, the effects of VNS are believed to be mediated by afferent stimulation and activation of the nucleus tractus solitarius and its central projections (Hachem et al., 2018). The most important of these projections are thought to be those to the locus coeruleus and the dorsal raphe nucleus. The locus coeruleus, which is activated rapidly in response to VNS, is the main noradrenergic nucleus of the central nervous system and projects to limbic structures (Hachem et al., 2018). The dorsal raphe nucleus modulates serotonergic pathways and appears to have a more delayed response to VNS.

More recently, novel approaches to non-invasive VNS stimulation (nVNS) have been developed. These use a handheld unit placed on the neck or stimulate the auricular branch of the vagus at the tragus of the external ear. The latter approach, transcutaneous auricular VNS (taVNS) has the advantage that it can be delivered using existing transcutaneous electrical nerve stimulation (TENS) devices. TENS is an established, affordable technology, available over the counter (or online) without prescription for self-administration without medical supervision. Our group has recently used this approach, with a clip electrode for attachment to the tragus, to show that using a TENS machine, we could enhance synaptic plasticity in a simple motor task in a sham-controlled study in healthy participants (McKeating et al., in preparation). Moreover, taVNS administered using a TENS device was used in the study investigating fatigue in patients with systemic lupus erythematosus described above (Aranow et al., 2021).

This mechanistic study will therefore investigate whether vagus nerve stimulation via the auricular branch (taVNS), self-administered using a TENS device, affects symptoms of fatigue and physiological, neurophysiological, behavioural, and immunological correlates of fatigue in otherwise healthy members of the public with pCF.

## Methods/Design

This protocol adheres to the SPIRIT guidelines (Chan et al., 2013) and a completed checklist can be found in the supplementary materials.

### Study design

The study is a single-site, single-blind, sham-controlled, randomized trial design, with all participants crossing over to the active intervention after 8 weeks. The study design is summarised in the Consort diagram in Figure 1.

**Figure 1:**
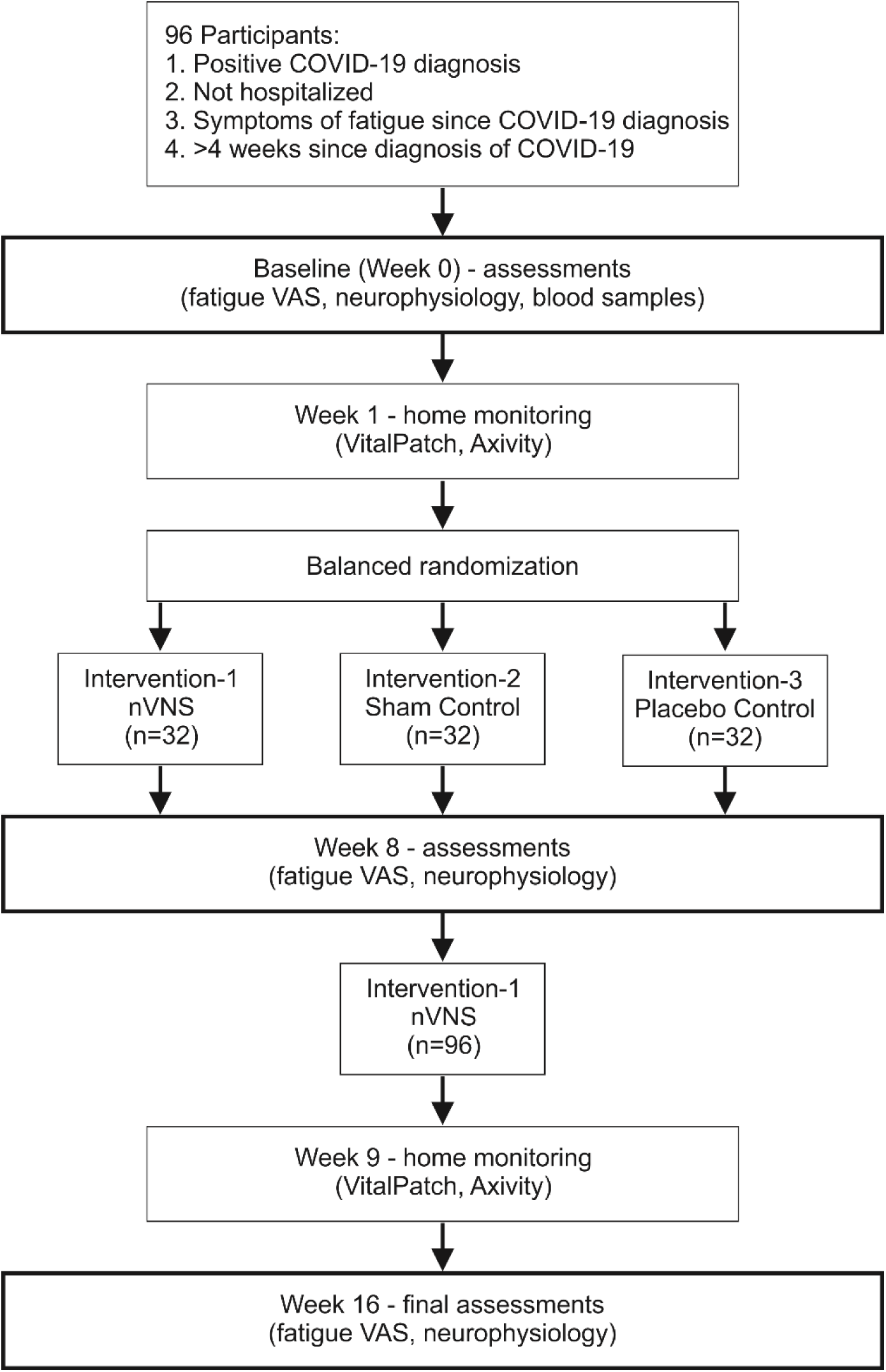
CONSORT diagram. VAS, visual analogue scale; nVNS, non-invasive vagus nerve stimulation. Participants randomized to the sham control group are not receiving any stimulation to the tragus due to a resistor shunting the current. Participants randomized to the placebo control group are receiving active stimulation to the earlobe.

Once eligibility is confirmed, consent will be taken, and baseline fatigue assessments and biometric, neurophysiological and behavioural assessments will be completed (week 0). Participants are fitted with wearable technology (VitalPatch Biosensor and Axivity wrist-worn accelerometer) and baseline ambulatory data is collected for 7 days. At baseline, participants are randomized to one of three experimental groups - active tragus stimulation, sham tragus stimulation (sham; control 1) and active pinna/greater auricular nerve stimulation (placebo; control 2).

Consumables and instructions for the assigned intervention are provided in a box.

At week 8, participants return to the lab and neurophysiological, behavioural and fatigue assessments will be repeated, and participants are provided with a new box of equipment, consumables, and instructions for the final phase of the intervention. Participants receiving active VNS will continue for a further 8 weeks (open-label phase of the study) and those initially in the control arms will crossover to the (open label) active intervention for the last 8 weeks of their participation in the study; giving all participants the opportunity to benefit from the active taVNS intervention. At week 16, all assessments are repeated in a final visit.

Assessments are done in the lab and one session takes 2 hours to complete.

Neither the study sponsor nor the funding body had any role in designing the study, or collecting, managing, or analysing the data. Modifications to the protocol will be updated in the trial registry and the updated versions will be made available to all relevant parties.

### Study setting

The study sponsor is Newcastle University, Kings Gate, Newcastle upon Tyne, NE1 7RU, UK, +44 (0)191 208 6000. The trial is carried out in the Henry Wellcome Building, Medical School, Newcastle University, Newcastle upon Tyne, UK.

As a single-site study, involving small numbers to test a clinical device with an established safety record, according to guidance from the funder (NIHR), neither a coordinating centre nor a data monitoring committee (DMC) were required, and therefore funding to support a coordinating centre or DMC was not requested. Moreover, a decision was made, in consultation with the funder, to merge the expert advisory committee and trial steering committee.

The study sponsor is permitted to inspect and audit the study at any time without providing notice.

### Participants and eligibility criteria

Members of the public between 18 and 65 years of age, who have a verifiable positive COVID diagnosis (polymerase chain reaction, lateral flow or antigen tests), but who did not require hospitalisation for the initial infection and who are at least 4 weeks after diagnosis will be recruited via the trial website (covidfatigue.co.uk) or through social media. They will be screened for symptoms of fatigue by answering two simple questions:

*Q1. How do your symptoms of fatigue affect your day-to-day living?*

Responses will be collected using a 0-10 Likert scale [0 = Not at all; 5 = Moderately; 10= Extremely].

*Q2. Do you feel that you need treatment for your fatigue: Yes or No*.

Only those who answer Yes to Q2 and score >/=3 in response to Q1 will be invited to take part.

Those with other medical issues that might affect fatigue assessment (previous diagnosis of neurological or psychiatric disorder, cardiac disease e.g., cardiomyopathy, myocardial infarction, arrhythmia, prolonged QT interval, etc.) are excluded. Participants are screened against additional contraindications of transcranial magnetic stimulation (implanted devices e.g. pacemaker, pregnancy). Participants have to be fluent in English in order to give written informed consent.Eligibility criteria are summarized in Table 1.

**Table 1:** Participant eligibility criteria.

| Inclusion Criteria | Exclusion Criteria |
| --- | --- |
| • 18 – 65 years of age<br>• Fluent in English<br>• Able to provide written informed consent | • Previous diagnosis of neurological or psychiatric disorder<br>• Cardiac disease (e.g. cardiomyopathy, myocardial infarction, arrhythmia, prolonged QT interval, etc.)<br>• Implanted device (e.g. pacemaker)<br>• Pregnant<br>• Contraindications for TMS (e.g. epilepsy, metal implants) |

Participants who meet eligibility criteria but are receiving medical therapy for a stable longterm condition should continue concomitant care. However, participants are advised that additional pharmacological or non-pharmacological (e.g., devices, behavioural therapies, etc.) interventions, which might exacerbate or improve symptoms of fatigue and thus interfere with study, are prohibited during the trial.

Volunteers are offered vouchers (£60 worth) for taking part in the trial.

### Randomization and blinding

Patients who provide informed consent to participate and fulfil the eligibility criteria are equally randomized to one of three groups (see Figure 1), one active and two control interventions, which they will self-administer for 8 weeks. Participants receiving active VNS will continue for a further 8 weeks and those initially in the control arms will crossover to the active intervention for the last 8 weeks of their participation in the study.

Balanced randomization will be performed independently by the statistical team using the random permuted blocks method. To ensure fairness an app will be used to display which group a patient has been placed in. Accesses to this information is logged to limit the IDs known to be in a specific group, which eliminates the possibility of a patient being deliberately assigned to a particular group.

Research team members need to view patient IDs and their respective group, however giving unrestricted access to this information could incur bias by allowing patients to be assigned to an ID that would place them inside a specific group. The app solves this by having user accounts with two different levels of access. The first is used by the research team members and only permits one patient’s group to be viewed at once and logs each one of these accesses. The second allows the statistical team to upload, edit and view all patient groups as well as view an audit log of all activity from research team members. It presents relevant options to the statistical team such as blocking or rate limiting a user from further access to patient group data.

The research associate (RA; or research team member) will set-up the stimulation parameters for the TENS device and train the participant to use the device at the end of their visit to the laboratory, after completing laboratory assessments. The RA will first explain to each participant that, ‘the aim of the study is to test the effects on fatigue of stimulating different parts of the ear lobe at different levels of stimulation’. The RA will then set stimulus parameters during the TENS demonstration by determining sensory threshold and pain/discomfort threshold for tragus, pinna and earlobe stimulation and instructing the participant to set the TENS at a specified current setting whenever it is used.

A small plastic junction box (see Figure 2Ac) connecting the TENS cables to the earclip cables contains either a 10 MΩ resistor for active stimulation or a 10 Ω resistor to shunt current. The resistor boxes are indistinguishable for participants. The RA is not blinded to the intervention after randomization.

**Figure 2:**
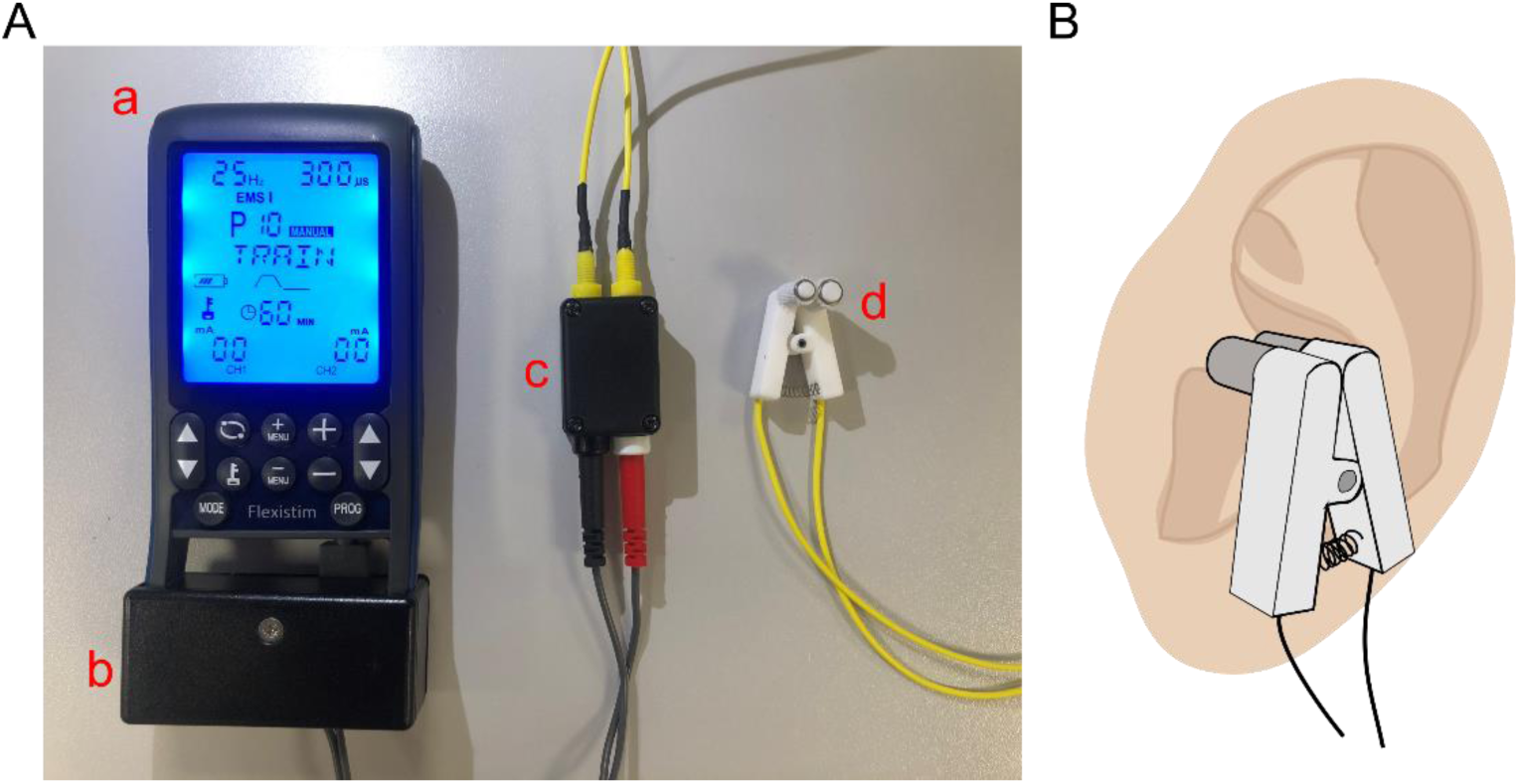
Transcutaneous electrical nerve stimulation (TENS) device. **A,** a, Flexistim TENS device; b, attached box containing data logger for monitoring usage; c, junction box containing either a 10 MΩ resistor for active stimulation or a 10 Ω resistor to shunt current; d, earclip with sleeves. **B,** Earclip positioning on tragus.

### Intervention

In this study, participants will be required to self-administer electrical stimulation to the external ear to activate nerves running beneath the surface of the skin. A FlexiStim (TensCare, Epsom, UK) transcutaneous electrical nerve stimulation (TENS) device will be used for this purpose (Figure 2Aa). These devices are commercially available over the counter, or on-line, without prescription, and having been specifically developed for safe use without training and with only minimal written instructions, participants should find these TENS devices straightforward to use.

To activate the auricular branch of the vagus nerve (taVNS) a clip electrode (Figure 2Ad & 2B) connected to the TENS system will be attached to the left tragus. Fabric sleeves, cut to the correct length to cover the two prongs of the ear clip electrode, ensure that the saline used to moisten the sleeves, makes a good electrical contact with skin overlying the tragus. To activate the greater auricular nerve (C2/C3 nerve roots; placebo control), the clip electrode will be attached to the left earlobe/pinna to stimulate this external ear region.

The stimulation parameters will be set for each participant using the electrical muscle stimulation (EMS) program on the TENS machine and the same parameters will be used for tragus and earlobe/pinna stimulation. The stimulation protocol will begin with a two-minute warm up at a frequency of 6Hz and pulse width of 200μs. This will be followed by a 60-minute train, which alternates 30 seconds at 25Hz and 300μs with 60 seconds at 4Hz and 200μs. Finally, there will be a 3-minute cool down stimulation protocol of 3Hz and 200μs. The stimulus current strength will be set just above perceptual threshold to ensure the subject is aware of the stimulation, but also so that the intensity is not uncomfortable or painful.

To ensure participants are blinded, for each intervention participants will only be required to power the TENS device ON (stimulator settings will be pre-set for each participant and the same settings used for all interventions). Instructions for delivering intervention-1, intervention-2 or intervention-3 (according to the randomization procedure) will be provided accordingly. The three interventions will be as follows:

*Intervention-1 (active)*: Clip electrode attached to the tragus and connected to the TENS device to stimulate the auricular branch of the vagus nerve i.e. Junction box connecting the TENS cables to the earclip cables contains a 10 MΩ resistor for active stimulation.

*Intervention-2 (sham control)*: Clip electrode, connected to a junction box containing a 10 Ω resistor to shunt stimulus current, is attached to the tragus such that the auricular nerve is not stimulated when the TENS device is powered on i.e. controlling for mechanical pressure exerted by the clip electrode on the tragus.

*Intervention-3 (placebo control):* Clip electrode attached to the earlobe and connected to the TENS device thus stimulating afferents of the greater auricular nerve (C2/C3 nerve roots) i.e. controlling for non-specific effects of electrical stimulation. Junction box connecting the TENS cables to the earclip cables contains a 10 MΩ resistor for active stimulation.

Before the participant starts the allocated intervention, the RA will also explain to each participant that, ‘because different current strengths are being tested, they may not feel the stimulation’.

Participants will be asked to apply the intervention three times per day.

Any adverse events (AE) will be diarised during the intervention and closely monitored until resolution, stabilisation or until it has been shown that the study procedures are not the cause.

### Adherence

In order to monitor adherence, a data logger is attached in a plastic box to the TENS device (see Figure 2Ab). The battery-powered data logger comprises a microcontroller, electrically erasable programmable read-only memory and a ferrite rod around which the TENS cable is looped. Every 10 minutes, the data logger powers up and determines if the TENS device is stimulating (measured as current passing through the TENS cable). Every 4 hours the data logger will write the count of ‘on’ states to the memory, which can then be used to determine how often the TENS device has been used over a given time period.

### Adverse Events

‘Adverse Event’ (AE) will be defined as any untoward medical event, including abnormal laboratory values, occurring during the use of the study device, but not necessarily causally related to it. Events with onset prior to start of study device use will not be considered as adverse events unless there is an increase in severity and/or frequency following institution of the study device.

Adverse events will include symptoms of which subjects complained during follow up visits as well as any signs detected by the subject’s physician or other health professional during routine medical appointments. Subjects will be instructed to contact the investigator immediately in the event of any physical discomfort while on the study. If a diagnosis is available, it will be noted as an AE; if no diagnosis is available each sign and symptom will be recorded as individual adverse events.

For each individual AE the nature of the event, duration, maximum intensity and finally subject outcome will be noted. Causality assessment will be done using the World Health Organization-Uppsala Monitoring Centre (WHO-UMC) Causality Assessment Criteria. AE severity will be graded as mild, moderate and severe. Subject-outcome criteria will be categorized as recovered, recovered with sequelae, AE persisting, died or data not available.

Subjects are free to withdraw from the study at any time, whether or not they experience an AE, without giving any reason and without their medical care or legal rights being affected.

### Assessment procedures

Table 2 illustrates a flow diagram summarising all study procedures and outcome measures. Assessments done in the lab can be completed in sessions lasting just under 2 hours.

**Table 2:**
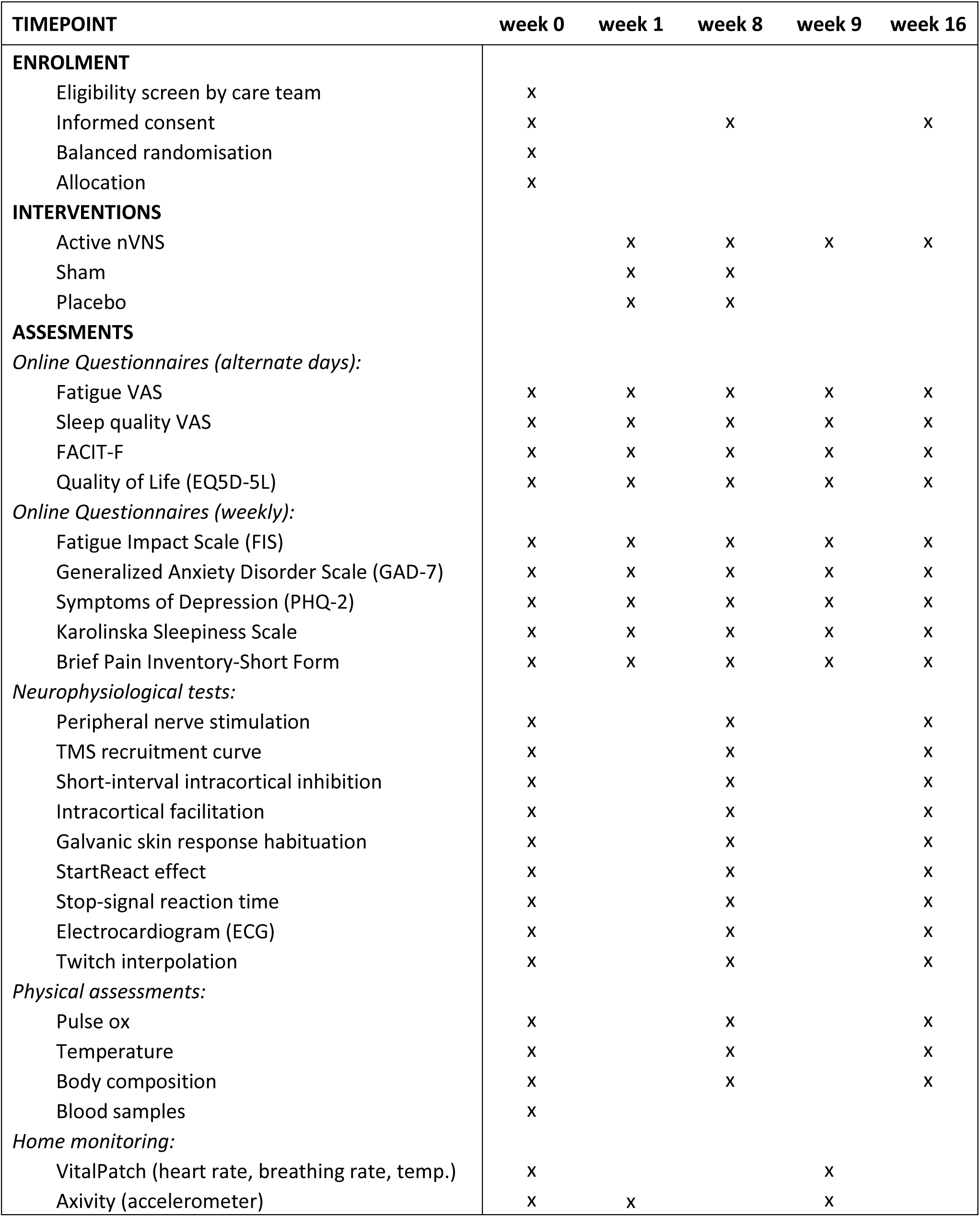
Flowchart of study procedure and outcome measures. Participants are required to visit the lab in week 0, week 8 and week 16 for assessments. Blood samples will be collected during one of three lab visits (preferably at week 0, but could be collected at week 8 or week 16).

### Questionnaires

Participants will be asked to fill out questionnaires online, over the course of the 16-week trial. Reminders will be sent out as text messages containing the link to fill out the correct questionnaire on their mobile.

The following short questionnaires will be delivered every other day: Fatigue Visual Analogue Scale (VAS), Sleep quality VAS, Functional Assessment of Chronic Illness Therapy Fatigue Scale (FACIT-F), Quality of Life (EQ5D-5L).

Once a week, a more detailed set of questions will be filled out including the following questionnaires: Fatigue Impact Scale (FIS), Generalized Anxiety Disorder Scale (GAD-7), Symptoms of Depression (PHQ-2), Karolinska Sleepiness Scale, Brief Pain Inventory-Short Form.

### Home monitoring

In addition to participant reported levels of fatigue, we will collect data on objective digital biomarkers of fatigue using CE marked wearables technology. This will involve a wrist-worn accelerometer (AX6, Axivity, UK) and a sticky patch on the chest (Vitalpatch, MediBioSense, UK; measures ECG, skin temperature, respiratory rate and energy expenditure among others). Accelerometer data will be collected for 14 days (week 0 & week 1) and 7 days (week 8). Vitalpatch data will be collected for 7 days during week 0 and week 8 respectively.

Android smartphones (Samsung Galaxy A32 5G Enterprise) will be given to participants, which will capture and store Vitalpatch data through the MediBioSense Health Mobile Application. The smartphones can also be used to answer the online questionnaires, should participants not own a smartphone or not wish to use their personal phone.

### General electrophysiological methods

The assessment protocols follow our previous study (Baker et al., 2023) which compared participants with pCF to age-matched controls. A summary of the various electrophysiological measures is illustrated in Figure 3.

**Figure 3:**
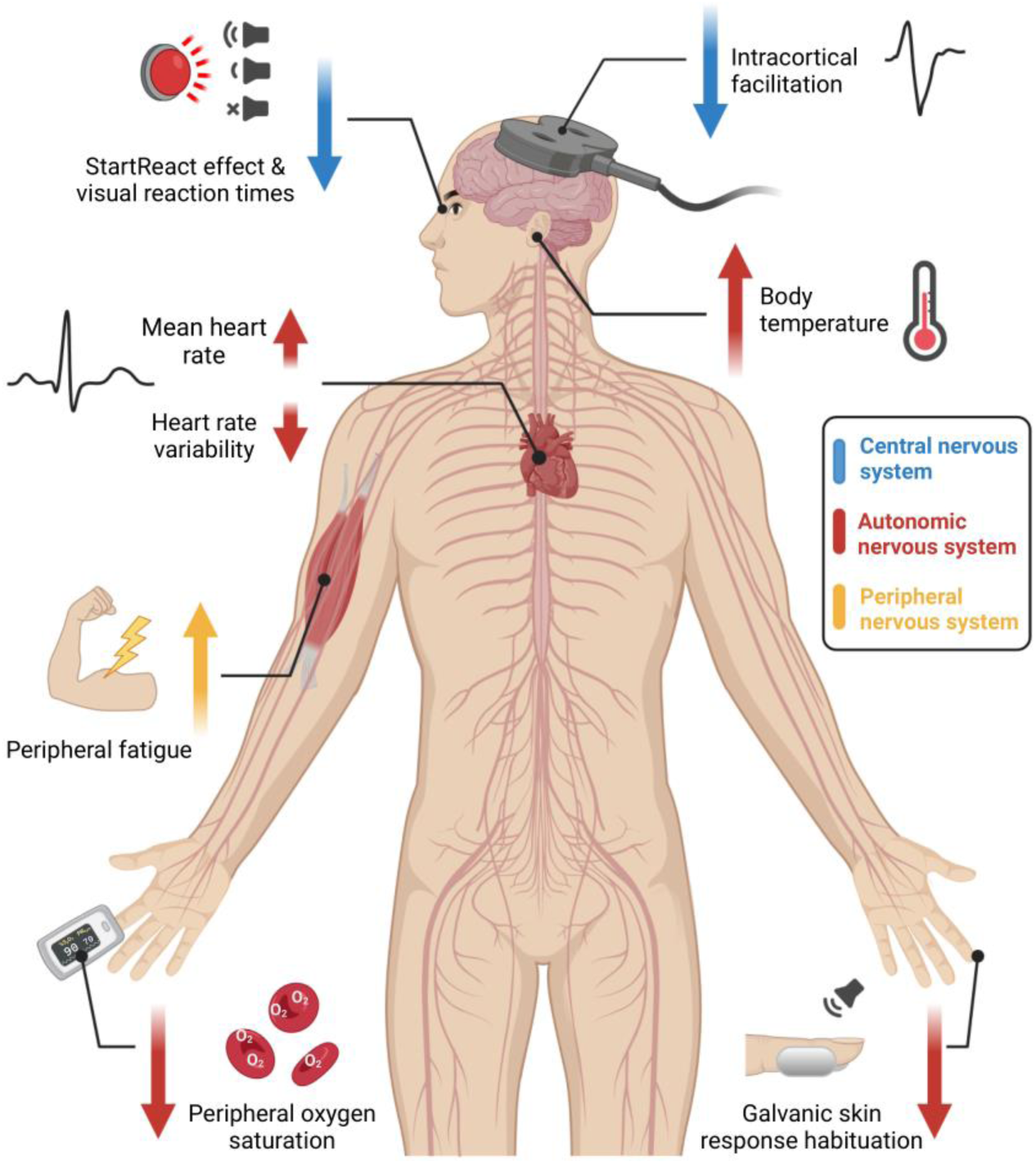
Schematic representation of neurophysiological assessments, colour coded according to which component of the nervous system (central; blue, peripheral; yellow, autonomic; red) they assess. Taken from (Baker et al., 2023).

Electromyogram (EMG) is recorded through adhesive surface electrodes placed on the skin over the muscle belly. EMG signals are amplified and filtered (band-width 30-2000 Hz) with a bioamplifier (D360 8-Channel Patient Amplifier, Digitimer, Welwyn Garden City, UK) and then converted to digital data with a sampling rate of 5kHz (CED Micro 1401 with Spike2 software, Cambridge Electronic Design, Cambridge, UK) and stored on a computer for off-line analysis. Where a measurement requires a constant contraction, visual feedback of rectified and smoothed EMG activity is provided to the subject via a display of coloured bars on a computer screen, calibrated to the subject’s maximum voluntary contraction (MVC). Participants are then asked to maintain a contraction corresponding to 10% of their individual MVC. Transcranial magnetic stimuli were applied using a figure-of-eight coil through a Bistim 200^2^ stimulator (Magstim, Whitland, UK). The magnetic coil is held to induce electrical currents that flow perpendicular to the presumed line of the central sulcus in a posterior-anterior direction, with the handle pointing backwards and 45° away from the midline. The hotspot was defined as the region where the largest motor-evoked potential (MEP) in the target muscle could be evoked. To ensure a stable coil position during experiments and across sessions, the site of stimulation is marked using a Brainsight neuronavigation system (Rogue Research Inc., Montréal, Canada), which allows online navigation. A Polaris Vicra camera (Northern Digital Inc., Canada) is used to track the coil.

### Peripheral nerve stimulation

Stimuli to peripheral nerves (0.2 ms pulse width) are delivered using a Digitimer DS7AH isolated, constant current stimulator. The size of the maximal M wave is measured by stimulating the median nerve at the wrist and recording EMG from the abductor pollicis brevis (APB) muscle. Stimulus intensity is set to produce a supra-maximal M wave. Ten stimuli at this intensity are delivered and the highest amplitude M wave will be used for subsequent normalisation of TMS recruitment curves (see below).

### TMS recruitment curve

As a measure of motor cortical excitability, the increase in APB MEP with increasing stimulus intensity will be used (Sangari and Perez, 2019). The active motor threshold (AMT) is defined as the intensity which produces a MEP > 100 µV amplitude in at least 3 out of 6 stimuli, while the participant maintains an active contraction of 10% MVC. The intensity of the stimulation was expressed as a percentage of the maximal stimulator output (MSO). Recruitment curves of increasing intensities in 10% MSO steps are obtained in blocks of ten stimuli per step starting at AMT intensity, until 100% MSO is reached.

Offline analysis will measure the size of MEPs from single trials and plot this versus stimulus intensity. A sigmoid curve can then be fitted to the relationship (Sangari et al., 2019).

### Paired-pulse TMS

The hotspot is defined as the region where the largest MEP in the first-dorsal interosseous (1DI) muscle can be evoked with the minimum intensity. Resting motor threshold (RMT) is defined as the minimal stimulus intensity needed to produce a MEP > 100 µV amplitude in at least 3 out of 6 stimuli.

For the test stimulus, TMS intensity is adjusted to elicit MEPs of 1mV amplitude at rest, or to 120% RMT, whichever is lower. The conditioning stimulus intensity is set at 80% RMT.

Conditioning stimulus precedes the test stimulus by 3 or 10 ms and the recorded responses are compared to responses to the test stimulus alone, to measure short-interval intracortical inhibition (SICI) and intracortical facilitation (ICF) respectively (Kujirai et al. (1993); see Figure 4A). 20 stimuli for each condition will be given in a pseudo-random order. Responses to the conditioning stimuli will be expressed as a percentage to the responses to the test stimulus alone.

**Figure 4:**
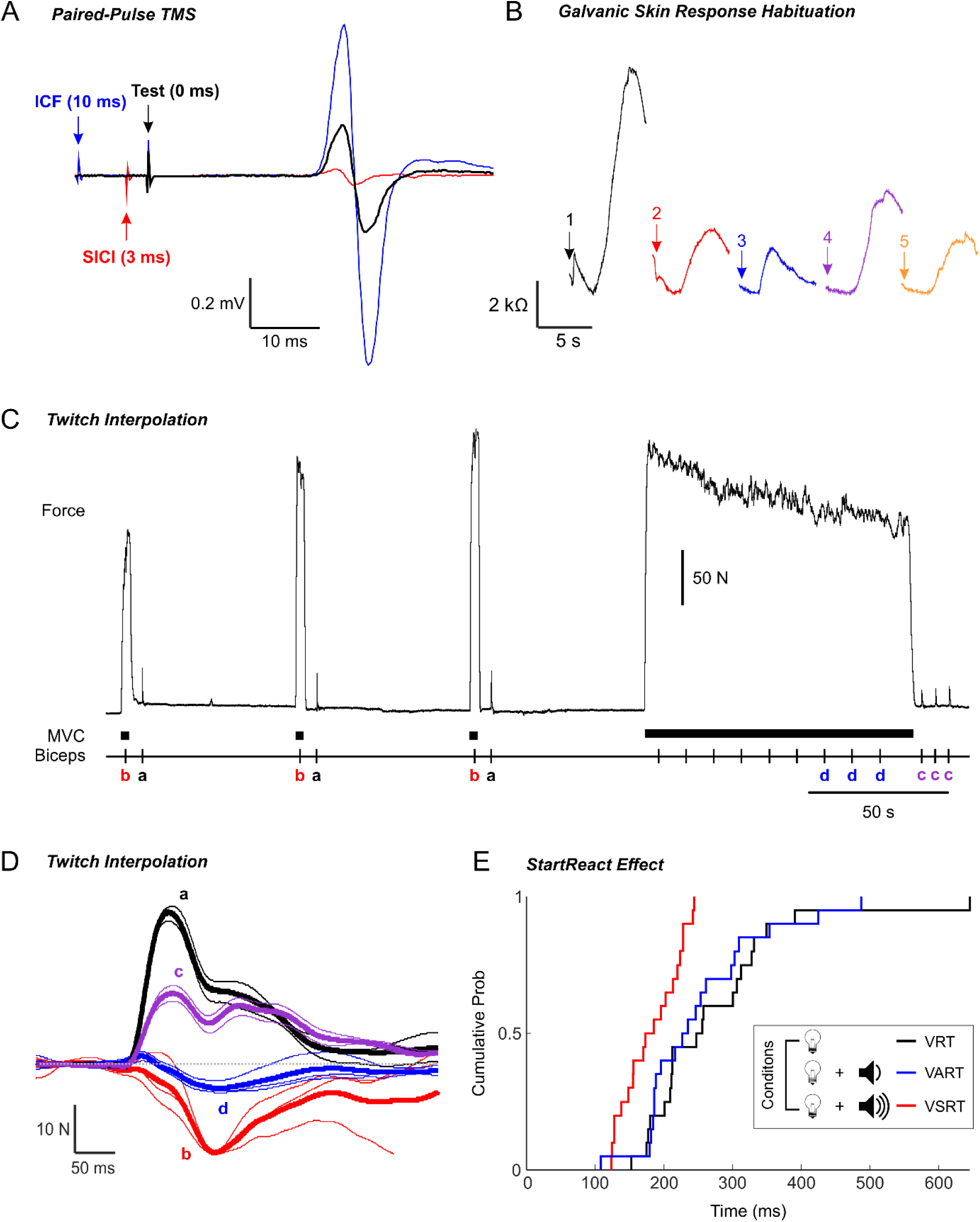
Single subject recordings exemplifying neurophysiological assessments. Taken from (Baker et al., 2023). **A**, Paired-pulse TMS: supra-threshold test stimulus preceded by a subthreshold conditioning stimulus at either 3 ms (short interval intracortical inhibition; SICI, red trace) or 10 ms (intracortical facilitation; ICF, blue trace) compared to test stimulus alone (black trace). **B**, Galvanic skin response habituation: change in skin conductance during five sequential loud sounds (arrows). **C**, Twitch interpolation: force trace and markers indicating times of MVC and delivery of electrical stimulation to the biceps muscle during the twitch interpolation paradigm. **D**, Average twitch amplitudes evoked by electrical stimulation to the biceps muscle under each condition as identified by the lower case bold letters in panel (C). **E**, StartReact: cumulative distribution of 1DI EMG onset times for all trials of a single subject when reacting to a visual cue alone (visual reaction time; VRT), a visual cue paired with a quiet auditory cue (visual auditory reaction time; VART), and a visual cue paired with a loud auditory cue (visual-startle reaction time; VSRT).

### Galvanic skin response

The galvanic skin response (GSR) is a measure of the cutaneous resistance or conductivity, which can be quantified by passing electricity through a pair of electrodes. Variation in skin resistance depends on sweat production, which itself is mediated by the sympathetic system (McCleary, 1950); its habituation may be a relevant measure in assessing cognitive states (Walker et al., 2019). The GSR is measured by placing two metal plates on the lateral and medial surfaces of the index finger. Five loud sounds (115 dB, C weighting, 500Hz, 50 ms, 6-6.8s inter-stimulus interval, chosen randomly from a uniform distribution) are played through loudspeakers placed underneath the subject chair, with the subject at rest. The ratio of the GSR response amplitude following the last stimulus compared to the first will be used as a measure of habituation (see Figure 4B).

### StartReact effect

This paradigm measures reaction time from EMG in response to a visual cue (visual reaction time, VRT), a visual plus quiet auditory cue (visual-auditory reaction time, VART), and a visual plus loud auditory cue (visual-startle reaction time, VSRT). The acceleration of reaction time between VART and VSRT is termed the StartReact effect and has previously been used to assess connections from the reticulospinal system (Fisher et al., 2013, Dean and Baker, 2017, Baker and Perez, 2017, Germann and Baker, 2021).

A green light-emitting diode (LED) is located ∼1 m in front of the subject. Participants are instructed to flex their elbow and clench their fist as quickly as possible after the LED illuminates. EMG is recorded from both the first dorsal interosseous muscle (1DI) and biceps muscle, and reaction time measured as the time from cue to onset of the EMG burst. Three types of trial were randomly interleaved (20 repeats per condition; inter-trial interval 6-6.8s, interval chosen randomly from a uniform distribution): LED illumination alone (VRT), LED paired with a quiet sound (80dB, 500Hz, 50ms, VART), LED paired with a loud sound (500 Hz, 50ms, 120 dB, VSRT).

StartReact measurements will be performed immediately after the GSR Habituation test, ensuring that any overt startle reflex has been habituated by the five loud sounds given in that test. The room lights will be dimmed for this test.

Data will be analysed offline trial-by-trial using a custom MATLAB program which identifies the reaction time as the point where the rectified EMG exceeded the mean + 7 SD of the baseline measured 0-200 ms prior to the stimulus. Every trial will also be inspected visually, and erroneous activity onset times (caused, for example, by electrical noise artefacts) will be manually corrected. Average VRT, VART and VSRT will be calculated for each subject and muscle, together with the amplitude of the StartReact effect, equal to VSRT-VART (see Figure 4E).

### Stop-signal reaction time

This stop-signal reaction time (SSRT) task measures the ability to inhibit a response after receiving a go cue. Participants are asked to respond to a go cue, but to inhibit their responses if a stop cue appears. This can indicate the state of premotor cortical-subcortical areas involved in impulse control (Logan and Cowan, 1984) and allows to test for increased levels of inhibition in the motor system. The hardware used to measure SSRT in this study is a portable device that was recently developed and uses Bayesian statistical analysis to improve the reliability of the measure (Choudhury et al., 2019). The battery-powered device consists of a plastic box which contains a microcontroller and a LCD screen, as well as one green LED, one red LED and a press button. Participants initiate a trial by pressing and holding down the response button. They are instructed to release this button as quickly as possible when the green LED illuminates (GO trial; 75% of trials).

On 25% of trials, the red LED illuminates 5, 65, 135 or 195ms after the green LED (stop trial) and participants are instructed not to release the button on these trials. Trials will be presented in three blocks of 64, with a 60 s break in between each block. Each block consists of 48 GO trials and 16 STOP trials (four for each delay). Using the distribution of reaction times on the GO trials, and the proportion of successfully inhibited responses, the algorithm calculates the SSRT as described in full in our previous work (Choudhury et al., 2019).

### Electrocardiogram

A single channel of electrocardiogram (ECG) recording will be captured, using a differential recording from left and right shoulder (bandpass 0.3-30 Hz) and stored for offline analysis. The time of each QRS complex will be extracted. From these times, the mean heart rate and the pNN50 (a measure of heart rate variability and defined as the proportion of successive intervals which differ by >50ms) are computed. ECG will be captured while participants are engaged in the SSRT test (see above), to ensure consistent behaviour across recordings.

### Twitch interpolation

Post-viral, immune-mediated disorders of the peripheral nervous system cause failure of signal propagation in nerve, ineffective signal transmission at the neuromuscular junction or reduced signal transmission in muscle fibres. If the same occurs in pCF, this would require a stronger voluntary drive to activate muscles and could give rise to a perception of effortful movements (de Vries et al., 2010). This will be tested before and after a maximal voluntary biceps muscle contraction by supramaximal electrical stimulation of the biceps muscle (twitch interpolation (TI); McDonald et al. (2010)).

Subjects sit with their arm and forearm strapped into a dynamometer to measure torque about the elbow. The forearm is held vertically in supination, the upper arm horizontal and the elbow is flexed at a 90° angle. Velcro straps hold the wrist, forearm and upper arm in place. Thin stainless-steel plate electrodes (30 x 15 mm) covered in saline-soaked gauze are used for electrical stimulation of the biceps muscle, by taping one electrode over the muscle belly and one over its distal tendon. The individual supramaximal stimulus level was set by increasing the intensity until the twitch response (recorded by the dynamometer) grows no further.

For a visual representation of the protocol see Figure 4CD. Upon receiving an auditory cue, subjects perform a 3s long maximal voluntary contraction until a stop tone sounds. Electrical stimulation to the biceps is given during MVC, 2s after the go cue and at rest, 5s after the stop cue. This sequence is repeated twice, with a 60 s rest period between go cues. After another 60s rest, another go cue then indicates to the subject to make a sustained MVC of up to 90s. If exerted force falls below 60% of the initial maximal level, contraction is terminated early. During this long MVC, biceps muscle is stimulated every 10s. A final three stimuli (inter-stimulus interval 5s) are given at rest.

If a subject truly performs a maximal voluntary contraction, a superimposed electrical stimulus should not be capable of generating extra force. The size of any elicited twitch thus measures a central activation deficit. Stimulation of a fatigued muscle at rest after the long contraction produces a smaller twitch than that seen before the sustained MVC, indicating peripheral fatigue.

Central activation before and after fatigue, as well as peripheral fatigue will be computed based on elicited twitches according to the calculations in McDonald et al. (2010).

### Biometric Data

Various biometric measurements will be collected from participants. These will include blood oxygen saturation, tympanic temperature, height, weight and full body composition (TANITA BC-545N Segmental Body Fat Scale).

### Blood sampling

Peripheral venous whole blood samples will be collected via a closed vacutainer system. Blood samples will be collected during one of three lab visits (week 0, week 8 or week 16). Blood samples will be stored within the Newcastle University Tissue Resource for analysis later in the project when sufficient samples are available for more labour and cost-effective batch runs to test for various molecular biomarkers (e.g. cytokines, adrenergic, cholinergic markers and transcriptomic biomarkers). Immune cell subset analysis will be performed by flow cytometry using fluorescent antibodies for T-cells, B-cells, NK-cells and monocyte subsets. Whole blood stimulation will be performed using 2ng/mL lipopolysaccharide (LPS) and compared to unstimulated whole blood. Blood will be stimulated for 24 hours in 48 well plates at 37°C and 5% CO2. The supernatants from each well will be harvested and the following cytokine levels measured by cytometric bead array: IFNγ, IL12-p70, TNFα, MIP-1α, IFNα, IL-10, IL-1β, IL-6 and IP10.

### Sample size

From data reported in (Tarn et al., 2019) and (Tarn et al., 2023), the fatigue VAS outcome had a pooled standard deviation of 25.1 at month 2, with a difference between arms of 17.4. As we are aiming to demonstrate the experimental arm is superior to both control arms, we aim for each comparison to have a two-sided 5% type I error rate and 88% power. This would give at least 80% conjunctive power (i.e. to show superiority to both controls).

Based on these figures, for a two-sided 5% error rate and 80% conjunctive power, we would need n=41 per arm. As we propose to use a linear regression with adjustment for baseline fatigue VAS, we will gain power if the baseline and follow-up measures are correlated. In data from Tarn et al. (2023), this correlation was 0.48. Using the formula from (Walters et al., 2019), we can reduce the sample size to 41*(1-0.48482) = 31.4 (rounded up to 32) per arm whilst maintaining the power at 88%.

### Analysis

The primary analysis will use a linear regression to estimate the difference in fatigue VAS between the active arm and each control arm as the primary outcome. The model will adjust for baseline fatigue VAS. An estimated treatment effect, confidence interval and p-value will be extracted from the model. To conclude the experimental arm is worthwhile, it will have to be superior to both control arms.

Due to short follow-up time, we anticipate minimal loss-to-follow-up so will utilise a complete cases analysis.

Due to adherence being important we have specified a complier average causal effect analysis to explore the anticipated treatment effect in a fully compliant population. Further details of this, and all other analyses, are provided in the Statistical Analysis Plan (SAP), provided in Supplementary File 1.

Data processing and statistical analyses will be conducted using MATLAB (TheMathWorks, Inc., Natick, MA, USA), SPSS (IBM SPSS Statistics for Windows, Armonk, NY: IBM Corp) and R (http://www.r-project.org/).

### Dissemination

The datasets generated during and/or analysed during the current study will be stored in a non-publicly available repository. Data will be uploaded to https://data.ncl.ac.uk. Data generated by this project will be in the form of computer files holding surface EMG signal waveforms recorded in response to stimuli. These files will be recorded by the Spike2 software package. Data will be made available after the researchers publish the papers arising from the study. Raw data is uploaded in anonymous form and participants will be asked to give consent for this at the beginning of the study. Participants will be sent the final paper, once published in a peer-reviewed journal.

## Discussion

Non-invasive vagus nerve stimulation has been shown to be an effective non-pharmacological treatment for primary headache, anxiety, depression and irritable bowel syndrome. There is now also evidence that nVNS can improve symptoms of fatigue associated with both primary Sjögren’s syndrome (Tarn et al., 2019, Tarn et al., 2023) and systemic lupus erythematosus (Aranow et al., 2021).

Fatigue is the most widely reported (Aiyegbusi et al., 2021) and potentially longest lasting (Stewart et al., 2022) symptom experienced by people suffering from long COVID. It’s devastating impact on quality of life calls for novel therapeutic options. Trans-auricular VNS has the advantage that it can be delivered using existing TENS devices, which are widely established, affordable and available over the counter.

This 16-week trial aims to investigate whether taVNS, self-administered using a TENS device at home, can reduce symptoms of fatigue in otherwise healthy members of the public experiencing pCF. The primary outcome measure is a fatigue visual analogue scale, which participants will give a score for every other day over the course of the trial.

We will use physiological, neurophysiological, behavioural, digital and immunological data to assess changes in the central, peripheral and autonomous nervous system that are hypothesized to mediate fatigue in pCF. Most of these measures have previously been shown to be significantly altered in pCF compared to healthy controls (Baker et al., 2023). Should taVNS improve symptoms of fatigue, we would expect these measures to normalise.

Secondary aims of the study are to:

1. Identify neurophysiological tests and blood biomarkers that might predict a response to nVNS and therefore permit treatment stratification in the future; and
2. Determine the optimum duration/dosing of nVNS therapy in pCF.

The data from the wearable monitoring devices used in Week 1 and Week 9 will allow us to determine when the effects of nVNS are first observed. Furthermore, by comparing the group differences at 8-weeks and 16 weeks we should be able to determine whether the effects of nVNS plateau or whether there are further incremental benefits of a more prolonged intervention.

Combining laboratory-based test of dysautonomia, cortical excitability and muscle pathology with subjective assessments and wearable technology, will allow this study to offer a detailed insight into the mechanisms responsible for post-covid fatigue.

## Data Availability

All data produced in the present work are contained in the manuscript

## Declarations

### Ethics approval and consent to participate

This study was approved by the Research Ethics Committee of the Faculty of Medical Sciences, Newcastle University (2284_1/18447/2021). This study is conducted in accordance with the Declaration of Helsinki. All participants are required to provide written informed consent by signing all study form(s), which are witnessed by an appropriately qualified and delegated member of the research team, prior to the completion of all study-specific procedures/ investigations.

### Consent for publication

Not applicable.

### Availability of data and materials

Not applicable. Data will be pseudo-anonymised using a unique identification code assigned at the first assessment and entered to a secure data system on Newcastle University computers for analysis.

### Authors’ contributions

M.G., A.S.B, J.M.S.W., M.S., S.C and M.R.B. wrote the main manuscript text

D.S.S. and S.N.B. prepared figures 3 and 4.

M.G. prepared tables 1 and 2, and figures 1 and 2.

M.G., F.N., D.S.S., S.N.B and M.R.B. conceived and designed the study

S.N.B. designed and contributed hardware to the study

A.S.B. designed and contributed software to the study

J.M.S.W., M.S. and S.C. were responsible for the statistical analysis plan

All authors reviewed the manuscript.

## Acknowledgements

Not applicable.

## List of abbreviations

1DI: First Dorsal Interosseous
AE: Adverse Event
AMT: Active Motor Threshold
APB: Abductor Pollicis Brevis
BMI: Body Mass Index
CFS: Chronic Fatigue Syndrome
COVID-19: Corona Virus Disease 2019
ECG: Electrocardiogram
EMG: Electromyogram
EMS: Electrical Muscle Stimulation
FACIT-F: Functional Assessment of Chronic Illness Therapy Fatigue Scale
FIS: Fatigue Impact Scale
GAD-7: Generalized Anxiety Disorder 7-item
GSR: Galvanic Skin Response
ICF: Intracortical Facilitation
MSO: Maximum Stimulator Output
MVC: Maximum Voluntary Contraction
nVNS: Non-invasive Vagus Nerve Stimulation
pCF: Post-Covid Fatigue
PHQ-2: Patient Health Questionnaire 2
RMT: Resting Motor Threshold
SARS-CoV-2: Severe Acute Respiratory Syndrome Coronavirus
SICI: Short-Interval Intracortical Inhibition
SSRT: Stop-Signal Reaction Time
taVNS: Transcutaneous Auricular Vagus Nerve Stimulation
TENS: Transcutaneous Electrical Nerve Stimulation
TI: Twitch Interpolation
TMS: Transcranial Magnetic Stimulation
VART: Visual-Auditory Reaction Time
VAS: Visual Analogue Scale
VNS: Vagus Nerve Stimulation
VRT: Visual Reaction Time
VSRT: Visual Startle Reaction Time

